# The Threshold for a Clinically Meaningful Improvement in Cardiopulmonary Exercise Testing Measures for Patients With Symptomatic Obstructive Hypertrophic Cardiomyopathy

**DOI:** 10.64898/2026.03.03.26347558

**Authors:** Ahmad Masri, Gregory D. Lewis, Roberto Barriales-Villa, Brian L. Claggett, Caroline J. Coats, Perry Elliott, Albert Hagège, Ian J. Kulac, Pablo Garcia-Pavia, Michael A. Fifer, Benjamin Meder, Iacopo Olivotto, Michael A. Nassif, Neal K. Lakdawala, Anjali T. Owens, Stephen B. Heitner, Daniel L. Jacoby, Regina Sohn, Stuart Kupfer, Fady I. Malik, Amy Wohltman, Martin S. Maron

## Abstract

**BACKGROUND:** Peak oxygen uptake (pVO_2_) is a strong, independent predictor of adverse cardiovascular outcomes, supporting cardiopulmonary exercise testing as a primary end point assessing efficacy of novel drug therapies in obstructive hypertrophic cardiomyopathy (oHCM) clinical trials. However, characterizing changes in pVO_2_ that patients perceive as beneficial or meaningful (ie, minimal important difference [MID]) has not been determined.

**METHODS:** Data from patients with symptomatic oHCM enrolled in SEQUOIA-HCM and MAPLE-HCM were pooled. A total of 282 patients were randomized 1:1 to aficamten (5–20 mg daily) or matching placebo in SEQUOIA-HCM, and 175 patients were randomized 1:1 to aficamten (5-20mg daily) or to metoprolol (50–200 mg) in MAPLE-HCM; follow-up in both trials was 24 weeks. Primary outcome was change from baseline to week 24 (Δ) in pVO_2_ using Patient Global Impression of Change with anchor-based analysis to define MID.

**RESULTS:** At week 24, ΔpVO_2_ (mL/kg/min) that corresponded to no change, one-category improvement, and one-category worsening were –0.05 (95% CI, –0.58 to 0.48), +0.35 (95% CI, –0.22 to 0.91), and –0.61 (95% CI, –1.36 to 0.13), respectively. Similarly, minute ventilation to carbon dioxide production ratio (VE/VCO_2_) slope that corresponded to no change, one-category improvement, and one-category worsening were 0.16 (95% CI, –0.59 to 0.90), –1.15 (95% CI, – 1.89 to –0.42), and 0.88 (95% CI, –0.42 to 2.19), respectively. In a responder analysis using this new threshold for pVO_2_, 60% of patients receiving aficamten achieved a ΔpVO_2_ ≥0.35 versus 31% of patients on placebo or metoprolol (odds ratio, 3.4 [95% CI, 2.3–4.9], *P*<0.001). Consistent findings were seen with VE/VCO_2_ responder analysis.

**CONCLUSIONS:** Changes in pVO_2_ of +0.35 and –0.61 mL/kg/min were associated with a small but perceptible clinical improvement and worsening, respectively, in patients with oHCM. Applying this newly defined threshold resulted in excellent differentiation of treatment effect in a clinical trial. These novel data provide a measure of clarity to patients and clinicians regarding the interpretation of changes in pVO_2_ following therapeutic interventions, with potential impact on HCM management strategies and future clinical trials.

**Clinical Trial Registration:** SEQUOIA-HCM (NCT05186818; https://clinicaltrials.gov/study/NCT05186818?term=sequoia-hcm&rank=1); MAPLE-HCM (NCT05767346; https://clinicaltrials.gov/study/NCT05767346?term=maple-hcm&rank=1)

**Clinical Perspective:** *What Is New?:*

- Using pooled data from over 440 patients with symptomatic obstructive hypertrophic cardiomyopathy enrolled in two phase 3 clinical trials, we define, for the first time, the minimally important difference for peak oxygen uptake (pVO_2_) and ventilatory efficiency (VE/VCO_2_) using patient-anchored and distribution-based methodologies.
- A change in pVO_2_ of +0.35 mL/kg/min and a change in VE/VCO_2_ of –1.15 represent the minimal thresholds associated with patient-perceived clinical improvement.
- Responder analyses using these thresholds demonstrated robust differentiation between aficamten and placebo/metoprolol, with an odds ratio exceeding 3 for achieving a meaningful improvement in pVO_2_.

*What Are the Clinical Implications?:*

- These newly defined thresholds bridge the gap between statistically significant changes in cardiopulmonary exercise testing measures and clinically meaningful benefit as perceived by patients with obstructive hypertrophic cardiomyopathy.
- Clinicians can use these benchmarks to contextualize individual patient responses to medical therapy, informing shared decision-making regarding treatment continuation or modification.
- These data provide a standardized, patient-centered framework for designing and interpreting primary end points in future hypertrophic cardiomyopathy clinical trials.

## INTRODUCTION

Cardiopulmonary exercise testing (CPET) provides an objective and reproducible assessment of functional capacity in patients with cardiovascular disease, with peak oxygen uptake (pVO_2_) representing a strong, independent predictor of adverse outcomes in obstructive hypertrophic cardiomyopathy (oHCM).^1-3^ In addition, pVO_2_ provides a comprehensive assessment of a number of pathophysiologic mechanisms in HCM that contribute to symptoms and functional limitation, including diastolic function and outflow tract obstruction. These particular strengths of CPET have supported its integration into clinical trials as a primary outcome measure in assessing efficacy of treatment interventions in HCM, including most recently cardiac myosin inhibitors. In this regard, statistically significant improvements in pVO_2_ with treatment have formed the basis for regulatory approval of these novel therapies for symptomatic oHCM.^4-6^

However, contextualizing the clinical relevance associated with changes in pVO_2_ to what patients might perceive as a meaningful alteration in symptom burden has not been defined in HCM. This is not necessarily a trivial point because treatment intervention can show statistically significant improvements without providing a meaningful benefit to patients. Therefore, given the increasing number of clinical trials with novel therapies in the HCM field, we felt it was timely to quantify the thresholds by which changes in pVO_2_ after a therapeutic intervention are perceived by patients, thus bridging the gap between statistical significance and patient-centered clinical meaningfulness.

To this end, using a multiparametric approach, we leveraged data from two recent oHCM clinical trials, SEQUOIA-HCM and MAPLE-HCM, to define the threshold in pVO_2_ (and other CPET variables) that is considered by patients to result in perceptible but important changes (ie, minimally important difference [MID]).^5^

## METHODS

### Study Design and Participants

SEQUOIA-HCM was an international phase 3 double-blind, randomized, placebo-controlled trial that evaluated the efficacy and safety of aficamten in addition to background medical therapy in 282 patients. The trial details have been previously described.^3,4^ MAPLE-HCM was an international phase 3, double-blind, double-dummy, head-to-head comparative trial in which patients with symptomatic oHCM received aficamten plus placebo (for metoprolol) or metoprolol plus placebo (for aficamten) in 175 patients. The trial details have been previously described.^5,7^ In addition to exercise capacity, measures of symptom burden,^8^ cardiac biomarker,^9^ and cardiac structure and function^10,11^ were assessed. Both studies followed an almost identical protocol, during which multiple and orthogonal metrics of efficacy were collected prospectively, allowing for high-fidelity assessment of treatment with either aficamten, placebo, or metoprolol in a longitudinal fashion. All participants provided informed consent, and the study was carried out in accordance with the provisions of the Declaration of Helsinki and the International Conference on Harmonization Good Clinical Practice guidelines.

Patients in both studies underwent symptom-limited CPET at baseline and at week 24 (end of treatment) and were evaluated in a blinded fashion by a core laboratory.^12^ The primary outcome was to define the MID for pVO_2_ as defined by the change from baseline to week 24 (Δ) in pVO_2_ in relationship to a 1-change of Patient Global Impression of Change (PGIC) at week 24. Secondary outcome measures were the MID for minute ventilation to carbon dioxide production ratio VE/VCO_2_ slope measured throughout exercise. PGIC was used as the primary anchoring measure, consistent with US Food and Drug Administration regulatory guidance.^13^ Briefly, the PGIC is a questionnaire that uses a 7-point scale to rate how patients consider their condition to have changed with treatment—1 (very much improved), 2 (much improved), 3 (minimally improved), 4 (no change), 5 (worse), 6 (much worse), and 7 (very much worse)—based on their overall personal assessment at different time periods throughout the study from baseline to week 24. Anchor-based MID was defined as the point change in the outcome of interest (pVO_2_, VE/VCO_2_, and CPET z-score) in patients reporting minimal improvement on the PGIC scale. Distribution-based MID was defined as the mean point change in the outcome of interest across the whole study population while accounting for the baseline value of the outcome of interest.

Secondary anchor measures included rest and Valsalva left ventricular outflow tract (LVOT) gradients and serum levels of cardiac biomarker N-terminal pro–B-type natriuretic peptide (NT-proBNP). Categories were derived based on thresholds deemed important by expert consensus. Predefined categorical absolute change from baseline to week 24 in LVOT gradients was used to derive five symmetrical categories: no change (±15 mmHg), improved (–15 to –45 mmHg), much improved (< –45 mmHg), worse (+15 to +45 mmHg), and much worse (> +45 mmHg). Proportional change from baseline to week 24 in NT-proBNP was used to define five categories of no change (±15%), improved (–15% to –45%), much improved (> –45%), worse (+15% to +45%), and much worse (> +45%). Using the defined MID thresholds for each outcome, a responder analysis was conducted with three categories: improved, no change, and worse. The primary statistical methodology for measures with known clinical relevance (PGIC) is anchor based, and for the secondary pathophysiologic metrics (LVOT gradient and NT-proBNP concentration) is distribution based. Both methods were conducted for each analysis for consistency.

### Statistical Analyses

Baseline characteristics were summarized using means and SDs for continuous normally distributed variables, medians and interquartile ranges for continuous non–normally distributed variables, and counts and percentages for categorical variables. Between-group comparisons were conducted using *t* tests, Wilcoxon rank-sum tests, and chi-squared tests, respectively.

Each anchoring variable considered in these analyses was either ordinal by nature (PGIC) or represents a categorized (ordinal) version of a continuous variable (eg, LVOT gradients, percent NT-proBNP change). For each ordinal anchoring variable and each continuous outcome, the mean value (and corresponding 95% CI) for each level of the anchoring variable is reported. For each outcome, the anchor-based thresholds for improvement and worsening are defined as the mean value observed amongst patients whose anchor values are one category above and below the designated “no change” level of the anchor variable. Next, the distribution-based thresholds were estimated using a linear regression model to quantify the expected change in the outcome variable per unit of the ordinal anchoring variable (accounting for the corresponding baseline value of the outcome variable).

MID thresholds based on these two methods were then used to perform responder analyses, where the MID thresholds were used to define no change, improvement, or worsening, in response to aficamten or placebo/metoprolol. Cumulative distribution function figures were used to visually represent the PGIC-anchored MID for PVO_2_ and VE/VCO_2_. All analyses were conducted using STATA (version 19, College Station, TX). *P* values <0.05 were considered statistically significant. No adjustments were made for multiple testing.

## RESULTS

### Baseline Characteristics

There were a total of 824 patients screened at 133 sites in 16 countries, and a total of 457 patients were randomized in both studies. The baseline characteristics for participants are shown in Table 1. Two hundred thirty patients were assigned to aficamten, and 227 patients were assigned to placebo or metoprolol. Of these, nine patients had missing PGIC data. Baseline characteristics were overall well balanced between groups, with a higher prevalence of hypertension in patients assigned to aficamten. Patients demonstrated substantially impaired exercise performance at baseline as assessed by CPET including pVO_2_ and VE/CO_2_. Table S1 provides a comparison between the SEQUOIA-HCM and MAPLE-HCM populations, showing well-balanced baseline characteristics and a protocol-driven lesser burden of disease in patients enrolled in the MAPLE-HCM study.

**Table 1.** Baseline Characteristics of Aficamten and Placebo/Metoprolol Groups.

| **Characteristic** | **Metoprolol +**<br>**placebo (n=227)** | **Aficamten**<br>**(n=230)** | ***P***<br>**value** |
| --- | --- | --- | --- |
| Age, y | 58.1 ± 13.3 | 59.1 ± 12.8 | 0.41 |
| Female sex, n (%) | 96 (42.3) | 92 (40.0) | 0.62 |
| Race, n (%) |  |  | 0.32 |
| White | 185 (81.5) | 178 (77.4) |  |
| Asian | 37 (16.3) | 42 (18.3) |  |
| Other | 5 (2.2) | 10 (4.3) |  |
| Geographic region, n (%) |  |  | 0.38 |
| Americas | 85 (37.4) | 94 (40.9) |  |
| China | 33 (14.5) | 35 (15.2) |  |
| Rest of world | 109 (48.0) | 101 (43.9) |  |
| Medical history |  |  |  |
| History of hypertension, n (%) | 103 (45.4) | 129 (56.1) | 0.022 |
| Known HCM-causing gene mutation, n (%) | 39 (17.2) | 38 (16.5) | 0.85 |
| Positive family history of HCM, n (%) | 52 (22.9) | 57 (24.8) | 0.64 |
| Paroxysmal atrial fibrillation, n (%) | 21 (9.3) | 21 (9.2) | 0.98 |
| Coronary artery disease, n (%) | 28 (12.3) | 27 (11.8) | 0.86 |
| Diabetes, n (%) | 19 (8.4) | 20 (8.7) | 0.90 |
| Permanent atrial fibrillation, n (%) | 1 (0.4) | 2 (0.9) | 0.57 |
| Implantable cardioverter defibrillator, n (%) | 30 (13.3) | 30 (13.1) | 0.96 |
| Vital signs |  |  |  |
| Systolic blood pressure, mmHg | 126 ± 15 | 125 ± 15 | 0.49 |
| Diastolic blood pressure, mmHg | 75 ± 10 | 76 ± 10 | 0.79 |
| Resting heart rate, beats/min | 74.4 ± 14.3 | 73.9 ± 13.3 | 0.68 |
| Body mass index, kg/m<sup>2</sup> | 28.2 ± 3.7 | 28.1 ± 3.6 | 0.71 |
| NYHA functional class, n (%) |  |  | 0.75 |
| II | 166 (73.1) | 171 (74.3) |  |
| III | 60 (26.4) | 59 (25.7) |  |
| IV | 1 (0.4) | 0 (0.0) |  |
| Biomarkers |  |  |  |
| NT-proBNP, pg/mL, median [IQR] | 546 [273, 1402] | 674 [304, 1330] | 0.45 |
| High-sensitivity troponin I, ng/L, median [IQR] | 12 [7, 25] | 13 [7, 34] | 0.17 |
| Cardiopulmonary exercise testing |  |  |  |
| Exercise modality, n (%) |  |  |  |
| Bicycle | 98 (43.2) | 100 (43.5) | 0.95 |
| Treadmill | 129 (56.8) | 130 (56.5) | 0.95 |
| pVO<sub>2</sub>, mL/kg/min | 19.2 ± 4.9 | 18.8 ± 4.6 | 0.38 |
| % predicted pVO<sub>2</sub> | 59.0 ± 13.3 | 57.6 ± 11.9 | 0.26 |
| VE/VCO<sub>2</sub> | 33.1 ± 5.8 | 33.4 ± 6.3 | 0.55 |
| Peak respiratory exchange ratio | 1.18 ± 0.10 | 1.18 ± 0.10 | 0.83 |
| Echocardiographic parameters |  |  |  |
| LVEF, % | 72 ± 7 | 72 ± 6 | 0.45 |
| Valsalva LVOT gradient, mmHg | 79 ± 33 | 80 ± 33 | 0.71 |
| Resting LVOT gradient, mmHg | 52 ± 31 | 52 ± 28 | 0.83 |
| Maximal LV wall thickness, mm | 20.93 ± 3.12 | 20.81 ± 2.93 | 0.67 |

Values are mean ± SD unless otherwise indicated.
HCM indicates hypertrophic cardiomyopathy; IQR, interquartile range; LV, left ventricular; LVEF, left ventricular ejection fraction; LVOT, left ventricular outflow tract; NT-proBNP, N-terminal pro-B-type natriuretic peptide; NYHA, New York Heart Association; pVO<sub>2</sub>, peak oxygen consumption; VE/VCO<sub>2</sub>, minute ventilation to carbon dioxide production ratio.

#### Relationship Between Exercise Parameters and Primary Anchor PGIC

At week 24, the distribution of the overall population across PGIC categories was as follows: 108 (24%) had no change, 120 (27%) minimally improved, 118 (26%) much improved, 67 (15%) very much improved, 28 (6%) worse, 6 (1.3%) much worse, and 1 (0.2%) very much worse (Table 2).

**Table 2.**
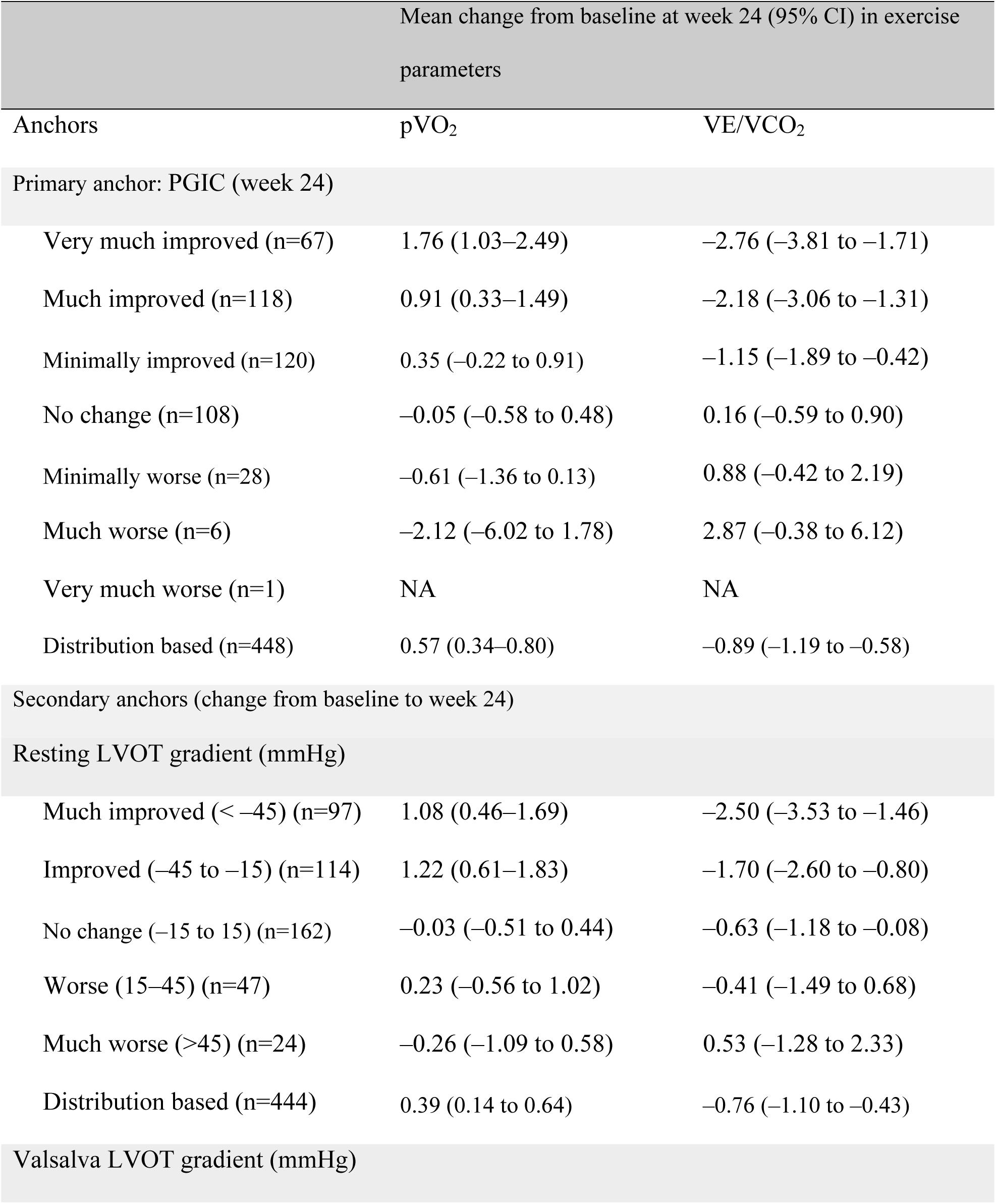
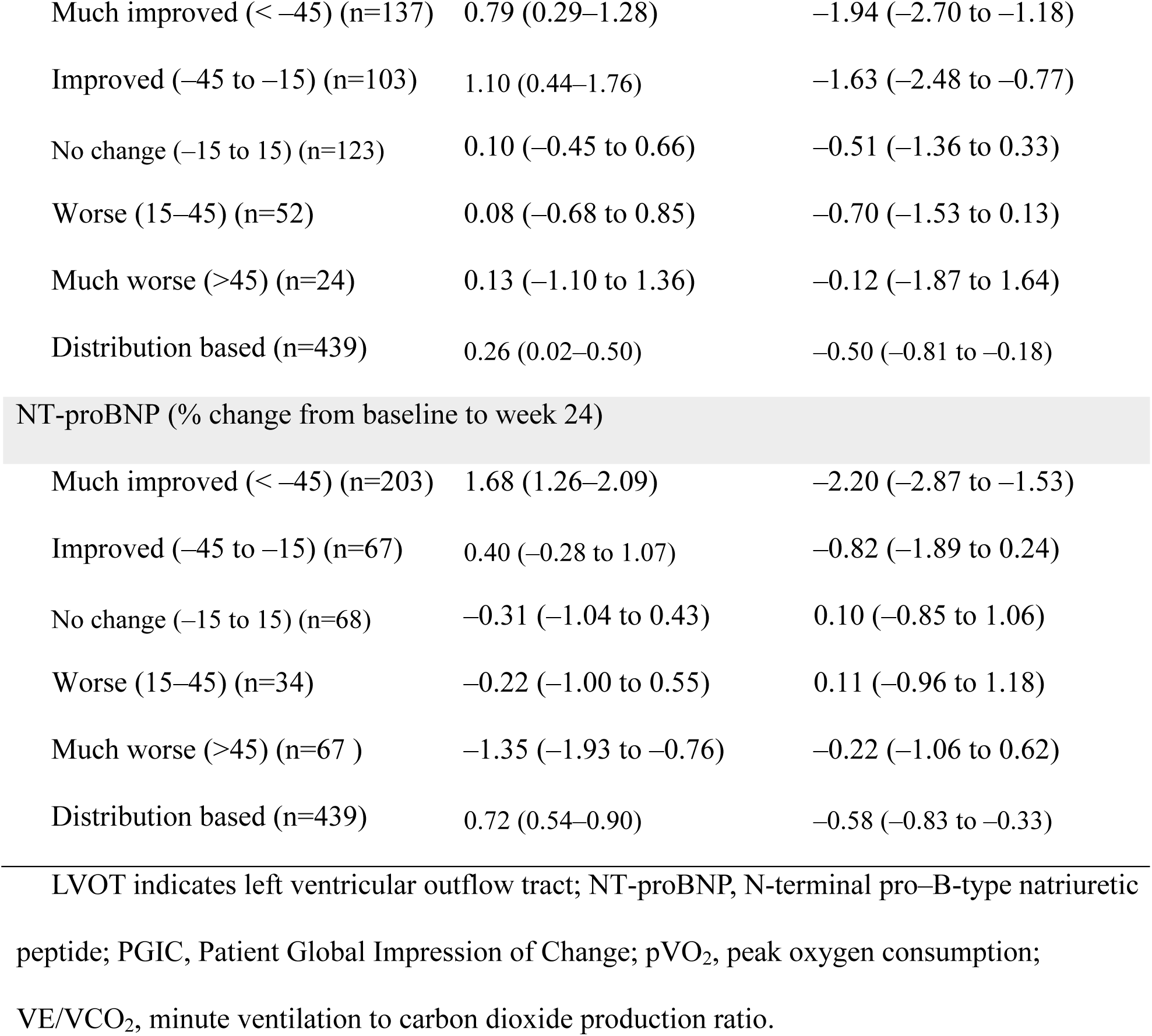
Mean Change in pVO_2_ and VE/VCO_2_ by PGIC Anchor and Disease Burden Distribution Analyses in SEQUOIA-HCM and MAPLE-HCM Trials.

|  | Mean change from baseline at week 24 (95% CI) in exercise parameters |  |
| --- | --- | --- |
| Anchors | pVO<sub>2</sub> | VE/VCO<sub>2</sub> |
| Primary anchor: PGIC (week 24) |  |  |
| Very much improved (n=67) | 1.76 (1.03–2.49) | –2.76 (–3.81 to –1.71) |
| Much improved (n=118) | 0.91 (0.33–1.49) | –2.18 (–3.06 to –1.31) |
| Minimally improved (n=120) | 0.35 (–0.22 to 0.91) | –1.15 (–1.89 to –0.42) |
| No change (n=108) | –0.05 (–0.58 to 0.48) | 0.16 (–0.59 to 0.90) |
| Minimally worse (n=28) | –0.61 (–1.36 to 0.13) | 0.88 (–0.42 to 2.19) |
| Much worse (n=6) | –2.12 (–6.02 to 1.78) | 2.87 (–0.38 to 6.12) |
| Very much worse (n=1) | NA | NA |
| Distribution based (n=448) | 0.57 (0.34–0.80) | –0.89 (–1.19 to –0.58) |
| Secondary anchors (change from baseline to week 24) |  |  |
| Resting LVOT gradient (mmHg) |  |  |
| Much improved (< –45) (n=97) | 1.08 (0.46–1.69) | –2.50 (–3.53 to –1.46) |
| Improved (–45 to –15) (n=114) | 1.22 (0.61–1.83) | –1.70 (–2.60 to –0.80) |
| No change (–15 to 15) (n=162) | –0.03 (–0.51 to 0.44) | –0.63 (–1.18 to –0.08) |
| Worse (15–45) (n=47) | 0.23 (–0.56 to 1.02) | –0.41 (–1.49 to 0.68) |
| Much worse (>45) (n=24) | –0.26 (–1.09 to 0.58) | 0.53 (–1.28 to 2.33) |
| Distribution based (n=444) | 0.39 (0.14 to 0.64) | –0.76 (–1.10 to –0.43) |
| Valsalva LVOT gradient (mmHg) |  |  |
| Much improved (< -45) (n=137) | 0.79 (0.29–1.28) | -1.94 (-2.70 to -1.18) |
| Improved (-45 to -15) (n=103) | 1.10 (0.44–1.76) | -1.63 (-2.48 to -0.77) |
| No change (-15 to 15) (n=123) | 0.10 (-0.45 to 0.66) | -0.51 (-1.36 to 0.33) |
| Worse (15–45) (n=52) | 0.08 (-0.68 to 0.85) | -0.70 (-1.53 to 0.13) |
| Much worse (>45) (n=24) | 0.13 (-1.10 to 1.36) | -0.12 (-1.87 to 1.64) |
| Distribution based (n=439) | 0.26 (0.02–0.50) | -0.50 (-0.81 to -0.18) |
| NT-proBNP (% change from baseline to week 24) |  |  |
| Much improved (< -45) (n=203) | 1.68 (1.26–2.09) | -2.20 (-2.87 to -1.53) |
| Improved (-45 to -15) (n=67) | 0.40 (-0.28 to 1.07) | -0.82 (-1.89 to 0.24) |
| No change (-15 to 15) (n=68) | -0.31 (-1.04 to 0.43) | 0.10 (-0.85 to 1.06) |
| Worse (15–45) (n=34) | -0.22 (-1.00 to 0.55) | 0.11 (-0.96 to 1.18) |
| Much worse (>45) (n=67 ) | -1.35 (-1.93 to -0.76) | -0.22 (-1.06 to 0.62) |
| Distribution based (n=439) | 0.72 (0.54–0.90) | -0.58 (-0.83 to -0.33) |

LVOT indicates left ventricular outflow tract; NT-proBNP, N-terminal pro-B-type natriuretic peptide; PGIC, Patient Global Impression of Change; pVO<sub>2</sub>, peak oxygen consumption; VE/VCO<sub>2</sub>, minute ventilation to carbon dioxide production ratio.

#### Peak Oxygen Uptake (pVO_2_)

Using PGIC as the primary anchor (Figure S1), the mean ΔpVO_2_ for no change, minimal improvement and minimal deterioration were –0.05 (95% CI, –0.58 to 0.48), +0.35 [95% CI, – 0.22 to 0.91) and –0.61 (95% CI, –1.36 to 0.13) respectively (Figure 1; Table 2). Distribution-based ΔpVO_2_ MID, including all 448 oHCM patients, was ±0.57 (95% CI, 0.34–0.80) (Figure S2). The between category differences, relative to no change, for minimal improvement and minimal worsening were +0.4 mL/kg/min and -0.56 mL/kg/min respectively.

**Figure 1.**
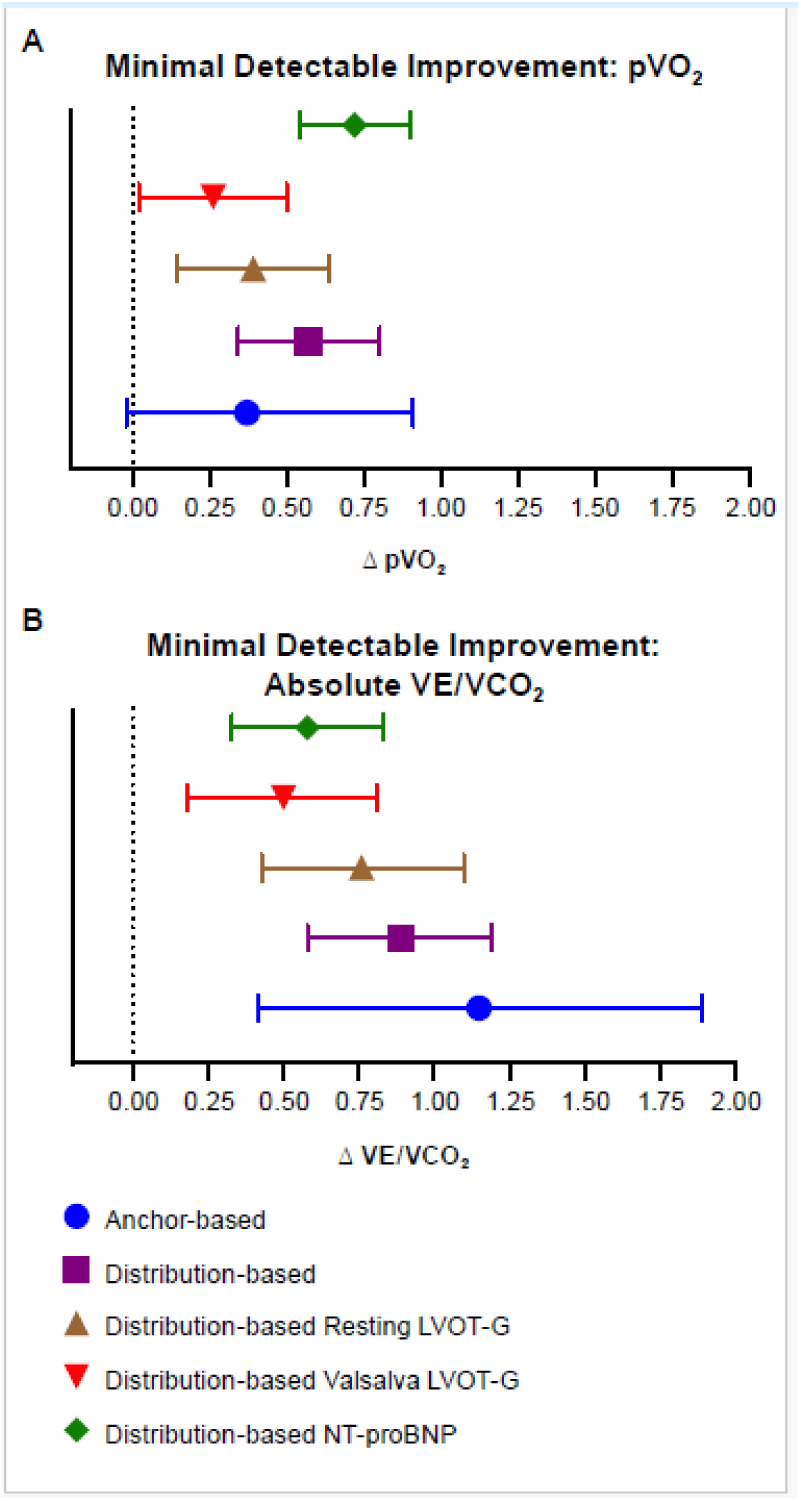
Minimal detectable improvement estimates for ΔpVO_2_ (A) and ΔVE/VCO_2_ (B). Points denote minimal detectable improvement values, and horizontal error bars indicate 95% CIs. Estimates are shown using anchor-based (blue) and distribution-based methods for resting LVOT-G (purple), Valsalva LVOT-G (red), and NT-proBNP (green), with the dotted vertical line marking no change (0). Δ indicates change from baseline; LVOT-G, left ventricular outflow tract gradient; NT-proBNP, N-terminal pro–B-type natriuretic peptide; pVO_2_, peak oxygen uptake; VE/VCO_2_, minute ventilation to carbon dioxide production ratio.

#### Submaximal Exercise (VE/VCO_2_)

Using PGIC as the primary anchor, the mean ΔVE/VCO_2_ for no change, minimal improvement and minimal deterioration were +0.16 (95% CI, –0.59 to 0.90), –1.15 (95% CI, –1.89 to –0.42), +0.88 (95% CI, –0.42 to 2.19) (Figure 1; Table 2). Distribution-based ΔVE/VCO_2_ MID, including all 448 oHCM patients, was –0.89 (95% CI, –1.19 to –0.58). The between category differences, relative to no change, for minimal improvement and minimal worsening were -1.31 and +0.72 respectively.

### Secondary Anchors

#### LVOT Gradients

The distribution of change in resting and Valsalva LVOT gradients is shown in Table 2. For resting LVOT gradients, 162 (37%) had no change in their gradient, 114 (26%) had improved gradient, 97 (22%) had much improved gradient, 47 (11%) had worse gradient, and 24 (5%) had much worse gradient. No change in resting LVOT gradient had a corresponding ΔpVO_2_ of –0.03 (95% CI, –0.51 to 0.44), and improved resting LVOT gradient yielded a ΔpVO_2_ of 1.22 (95% CI, 0.61–1.83). For Valsalva LVOT gradient, 123 (28%) had no change in their gradient, 103 (24%) had improved gradient, 137 (31%) had much improved gradient, 52 (12%) had worse gradient, and 24 (6%) had much worse gradient. No change in Valsalva LVOT gradient corresponded with a ΔpVO_2_ of 0.1 (95% CI, –0.45 to 0.66), and reduction of Valsalva LVOT gradient corresponded with a ΔpVO_2_ of 1.1 (95% CI, 0.44–1.76). The distribution-based approach generated ΔpVO_2_ MIDs of 0.39 (95% CI, 0.14–0.64) and 0.26 (95% CI, 0.02–0.5) for resting and Valsalva LVOT gradients, respectively. The results for VE/VCO_2_ followed a similar trend (Figure 1; Table 2).

#### NT-proBNP

The distribution of change in NT-proBNP is shown in Table 2. Sixty-eight (15.5%) had no change, 67 (15%) had improved levels, 203 (46%) had much improved levels, 34 (8%) had worse levels, and 67 (15%) had much worse levels. No change in NT-proBNP corresponded with a ΔpVO_2_ of –0.31 (95% CI, –1.04 to 0.43), and improvement in NT-proBNP corresponded with a ΔpVO_2_ MID of 0.4 (95% CI, –0.28 to 1.07) for anchor-based and 0.72 (95% CI, 0.54–0.90) for distribution-based approaches. The results for VE/VCO_2_ followed a similar trend (Figure 1; Table 2). In addition, Figure 2 provides visual evidence of the distribution of change in each outcome, using cumulative distribution curves of patients receiving aficamten or placebo/metoprolol.

**Figure 2.**
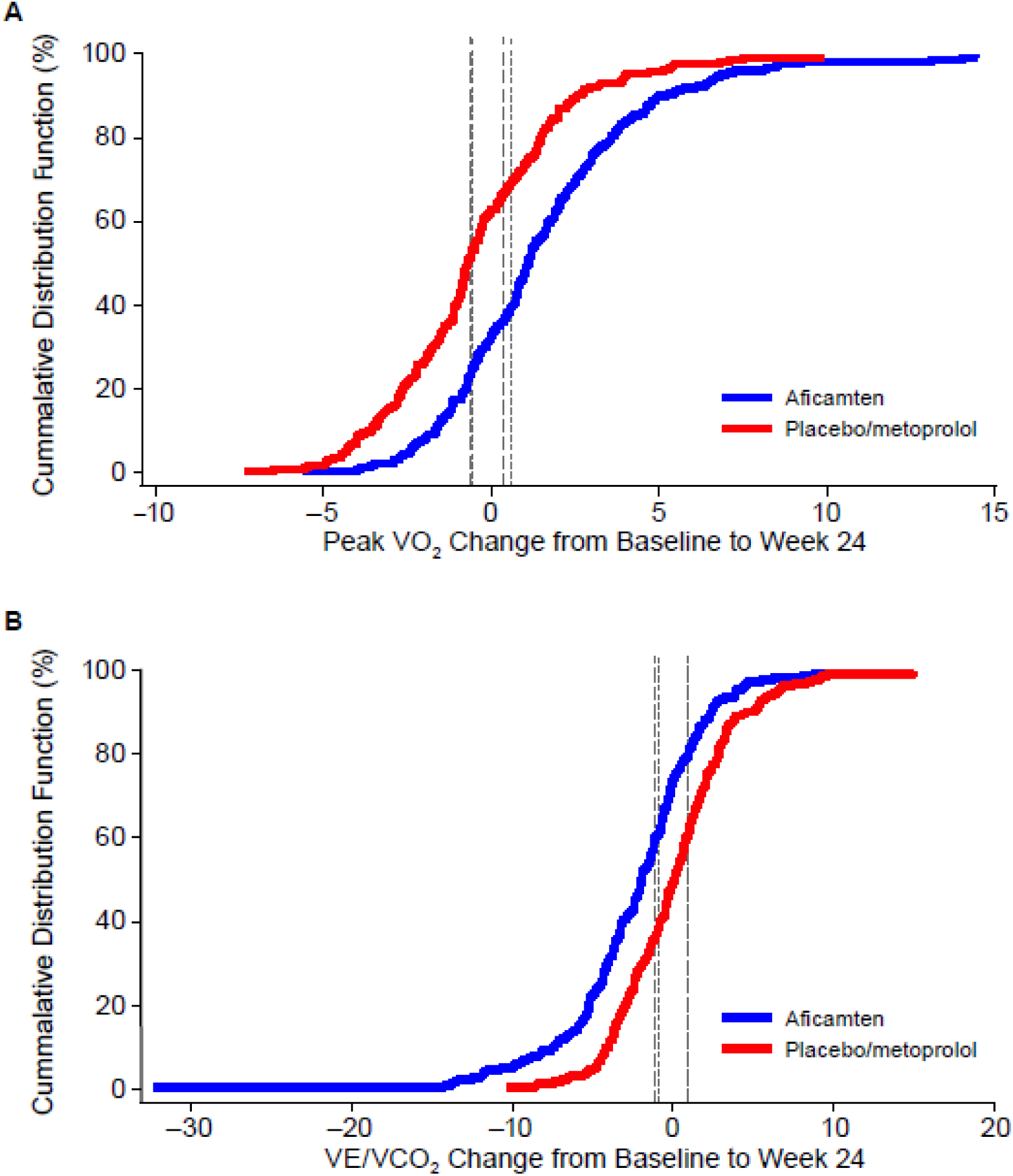
Cumulative distribution functions for change from baseline to week 24 in ΔpVO_2_ (A) and ΔVE/VCO_2_ (B), comparing aficamten (blue) versus placebo/metoprolol (red). Vertical dashed lines denote the MID used to classify response with 95% CI. Δ indicates change from baseline; MID, minimal important difference; pVO_2_, peak oxygen uptake; VE/VCO_2_, minute ventilation to carbon dioxide production ratio.

### Responder Analysis

The anchor- and distribution-based results of responder analysis for all outcomes are shown in Figure 3 and Table S2. Applying the anchor-based MID, 138 (60%) of 230 patients receiving aficamten achieved a ΔpVO_2_ ≥0.35 mL/kg/min versus 70 (31%) of 227 patients receiving placebo or metoprolol (odds ratio [OR], 3.4 [95% CI, 2.3–4.9], *P*<0.001). A worsening in exercise capacity with ΔpVO_2_ ≤-0.61 mL/kg/min was observed in a greater proportion of patients assigned to placebo/metoprolol versus aficamten (48% vs 22%; OR, 3.2 [95% CI, 2.2–4.9], *P*<0.001). The responses by treatment arm in each study (SEQUOIA-HCM and MAPLE-HCM) were similar (Table S3), and as such the responder analysis was applied to the aficamten group versus the placebo or metoprolol group subsequently.

**Figure 3.**
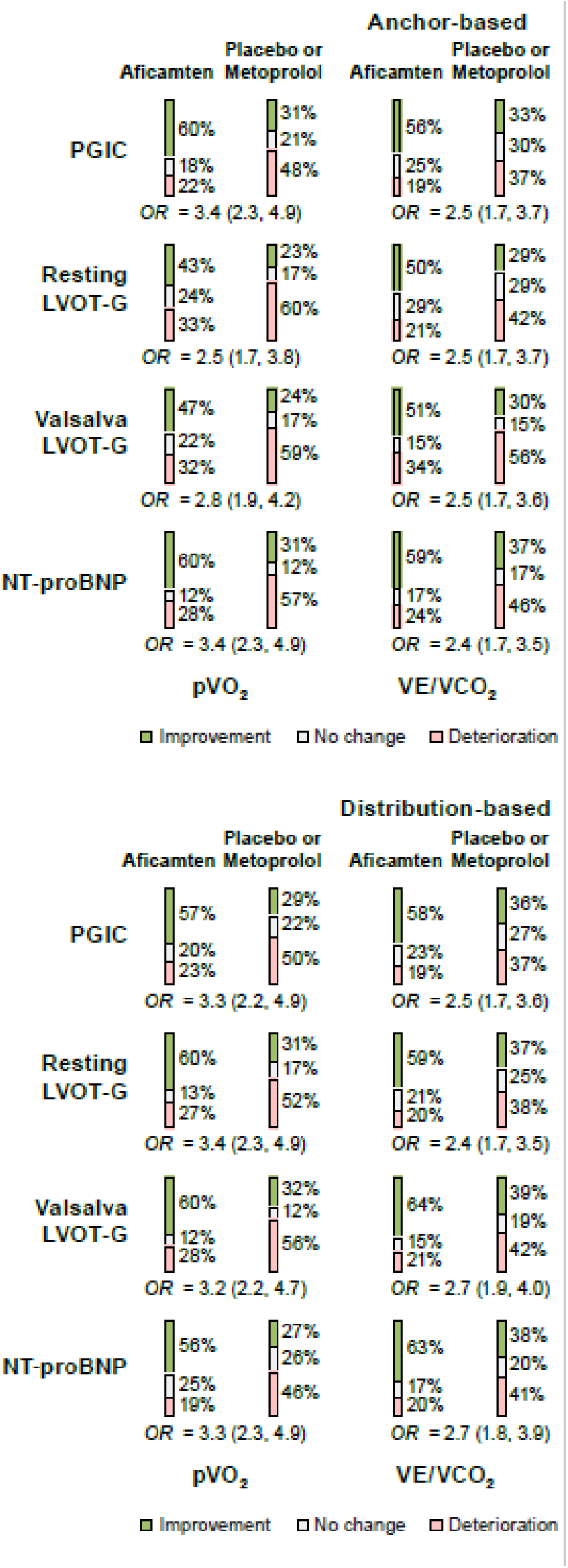
Responder analysis for each anchor and outcome using anchor- and distribution-defined MID. Proportions of participants classified as improved, unchanged, or deteriorated for each exercise end point using anchor-based (top) and distribution-based (bottom) thresholds, comparing aficamten versus placebo or metoprolol. Classifications are shown for ΔpVO_2_ and ΔVE/VCO_2_, using anchors of PGIC, resting LVOT-G, Valsalva LVOT-G, and NT-proBNP. ORs with 95% CIs are displayed for the likelihood of improvement with aficamten versus placebo or metoprolol. Δ indicates change from baseline; LVOT-G, left ventricular outflow tract gradient; MID, minimal important difference; NT-proBNP, N-terminal pro–B-type natriuretic peptide; OR, odds ratio; PGIC, Patient Global Impression of Change; pVO_2_, peak oxygen uptake; VE/VCO_2_, minute ventilation to carbon dioxide production ratio.

Applying the distribution-based MID, 131 (57%) of 230 patients on aficamten experienced a ΔpVO_2_ ≥0.57 mL/kg/min versus 65 (29%) of 227 patients receiving placebo or metoprolol (OR, 3.3 [95% CI, 2.2–4.9], *P*<0.001). Independent of the analytical strategy, the proportions of those improving, worsening, or remaining unchanged were fairly similar between groups (Figure 3; Table S2).

## DISCUSSION

Over the past several decades, efficacy of therapeutic interventions in HCM have been evaluated primarily based on changes in pVO_2_. This precedent is based on the principle that CPET provides objective measures of improvement in exertional capacity (an important treatment goal in HCM), and, as a powerful prognostic marker, significant improvements in pVO_2_ provide support that the treatment intervention is likely to beneficially alter the natural history of HCM patients. Recently, results from phase 3 clinical trials in oHCM have supported regulatory approval of cardiac myosin inhibitor drugs by demonstrating statistically significant improvements in pVO_2_ with therapy compared to placebo.^4-6,14^ However, the clinical relevance to HCM patients associated with statistically significant improvements in pVO_2_ observed in clinical trials has not been well defined. This knowledge gap has made it more challenging to completely interpret changes in pVO_2_ associated with drug therapy in oHCM. This has also potentially limited our understanding of the benefit HCM patients may expect to experience from a given treatment. Therefore, a framework to specifically contextualize the clinical meaning associated with pVO_2_ results, including relationship to the patient-centric treatment goal of improving patient well-being or other relevant disease-related measures, is a current unmet need in this disease.

To this end, we leveraged comprehensive and detailed outcome data from more than 440 prospectively followed symptomatic oHCM patients in two contemporary phase 3 clinical trials. To determine the MID for pVO_2_ in oHCM, we employed contemporary methodology previously recommended by regulatory agencies. With the anchor-based analytic strategies, we identified a change in pVO_2_ of +0.35 mL/kg/min to represent the minimal change in exercise capacity that is associated with a perceived benefit by patients, and a change of –0.61 mL/kg/min to be associated with a corresponding worsening in clinical status. Additionally, we assessed the change in maximal and submaximal exercise parameters using different analytic methodologies and pathophysiologic metrics to fortify the scientific robustness of the MID for pVO_2_ and VE/VCO_2_. While these pathophysiologic measures demonstrated excellent discriminatory capability as seen by separation on their respective cumulative distribution functions, they lack a foundation in clinical relevance (ie, patients do not “feel” their obstruction or NT-proBNP concentration). Additionally, there are no known thresholds of clinical meaningfulness for changes in LVOT gradient or NT-proBNP, further supporting the implementation of distribution-based methodology. Thus, an MID of 0.4–0.6 mL/kg/m^2^ is consistent with the values generated using measures of LVOT gradients (rest and Valsalva LVOT gradient MID of 0.4 and 0.3 mL/kg/m^2^, respectively) and NT-proBNP concentration (MID of 0.7 mL/kg/m^2^).

Historically, evidence from the heart failure literature^15^ and observational data in HCM^1,2^ have demonstrated that for every ∼1 mL/kg/min increase in pVO_2_, the relative risk of death and transplant decreases 21%, defining what is considered to be a clinically relevant difference with CPET and the efficacy bar to which statistically significant changes in pVO_2_ from prior HCM clinical trials have been considered. However, the threshold of 1 mL/kg/min was not anchored to patient perception nor pathophysiology and does not provide insight into what magnitude of change in pVO_2_ represents a perceived clinical benefit to an individual HCM patient. Our currently defined MID for pVO_2_ is also lower than the threshold used to define an exercise responder in the primary end point of EXPLORER-HCM, and subsequently in exploratory analyses from both SEQUOIA-HCM and MAPLE-HCM (1.5 mL/kg/min)^1,2,4,6,16^; however, this threshold appears to have been empirically chosen. It stands to reason that the currently derived lower threshold (pVO_2_ ∼0.5 mL/kg/min), the minimal perceptible difference, would be lower than that which is associated with significant morbidity and mortality (pVO_2_ ∼1.0 mL/kg/min). Additionally, because the treatment goals for patients with oHCM are to primarily improve functional capacity and quality of life, the current analysis aligns with this aspiration by defining, for the first time, a threshold targeting this outcome. Specifically, one might consider defining the treatment effect as the fold-increase in the MID of pVO_2_ (eg, treatment with aficamten led to a five-fold MID increase relative to placebo in SEQUOIA-HCM), which is clinically interpretable by both physicians and their patients.

We conducted a responder analysis as a demonstration for the implementation of the MID for pVO_2_ and VE/VCO_2_. Here we show that the OR for a response was >3 using either of the CPET measures, with the majority of aficamten-treated patients achieving this threshold. It is also important to acknowledge that there were patients who did not improve their exercise capacity in response to treatment; however, there are naturally multiple, often independent, parameters by which treatment success can be achieved. As such, one should be careful applying these thresholds for clinical decision making in isolation with individual patients without integrating the overall clinical context (eg, a patient who might feel much improved with reduction in LVOT gradients, but with improvement in pVO_2_ below 0.5 mL/kg/min, is still benefiting and should likely continue their treatment).^17,18^

Our findings carry major implications for interpretation of future clinical trials, putting forth a robust patient-centered threshold that holds clinical meaning beyond analysis for statistical differences. Furthermore, while traditional approaches for defining MID for clinical trial end points have relied solely on subjective anchors, we further fortify our measure using orthogonal measures of disease burden in an effort to more precisely triangulate the MID. However, we acknowledge that a substantial limitation remains. By comparison to other disease states, the relatively small population and short duration of the studies used in this analysis, and the low rate of major adverse cardiovascular events rates in the oHCM population, we are unable to effectively evaluate the MIDs through the lens of hard clinical end points. Nonetheless, the primary focus of generating these thresholds was to enable the placement of CPET data generated from trials into clinical context and is ultimately best defined as a meaningful change by patients themselves.

In conclusion, by leveraging unique comprehensive clinical trial data from a large cohort of oHCM patients, we have expanded our understanding regarding the clinical relevance associated with changes in pVO_2_ and VE/VCO_2_. Improvements of approximately +0.35 mL/kg/min in pVO_2_ and –1.15 in VE/VCO_2_ represent the minimal thresholds for improvement in these CPET-derived measures that is meaningful as perceived by patients, Therefore, these data bridge the gap between statistical significance and clinical meaningfulness. We believe that this knowledge will ultimately help patients and clinicians more clearly contextualize and appreciate the relevance of CPET-based responses to treatment interventions and should be considered when implementing and interpreting results of future HCM clinical trials.

## Non-standard Abbreviations and Acronyms

CPET: cardiopulmonary exercise testing
HCM: hypertrophic cardiomyopathy
LV: left ventricular
LVEF: left ventricular ejection fraction
LVOT: left ventricular outflow tract
LVOT-G: left ventricular outflow tract gradient
MID: minimally important difference
NT-proBNP: N-terminal pro–brain natriuretic peptide
NYHA: New York Heart Association
oHCM: obstructive hypertrophic cardiomyopathy
OR: odds ratio
PGIC: Patient Global Impression of Change
pVO_2_: peak oxygen uptake
VE/VCO_2_: ventilatory efficiency

## Acknowledgments

We thank the following individuals for their contributions to this clinical trial: participants and their families; investigators and study site staff; and Data Monitoring Committee members.

## Data availability

Qualified researchers may submit a request containing the research objectives, endpoints/outcomes of interest, Statistical Analysis Plan, data requirements, publication plan, and qualifications of the researcher(s). Requests are reviewed by a committee of internal and external advisers. If approved, data necessary to address the research question will be provided under the terms of the data sharing agreement. Requests will be considered after applications for marketing authorization in the US and Europe have been reviewed and final decisions rendered. Requests may be submitted to.

## Supplemental Material

Figures S1–S2 Tables S1–S2

